# Cancer-Tissue Fraction as a Scanner-Robust Triage Signal for Automated Gleason Grading of Prostate Biopsies: External Validation Across a Middle Eastern Cohort

**DOI:** 10.64898/2026.07.28.26359127

**Authors:** Joshua L Ebbert, Anthony Perry, Jared Szymanski, Dennis Della Corte

## Abstract

**Background:** Deep-learning systems for Gleason grading are developed almost entirely on high-end clinical scanners and on cohorts from a small number of Western institutions, yet deployment increasingly involves other devices and other populations. These two distribution shifts, device and population, are rarely tested together on the same physical slides. The PAR dataset, from Erbil, Iraq, digitizes each biopsy on three scanners and provides three distinct pathologist grades, so it permits both tests at once on a Middle Eastern cohort. A concurrent study by the dataset originators validated a task-specific model and two foundation models on PAR; we complement it by testing an independently developed detect-then-grade pipeline and by separating scanner effects on detection from scanner effects on grading.

**Methods:** We applied one fixed model developed on North American and European material to all 1017 whole-slide images (339 slides from 185 patients, three scanners; 49.6% clinically significant cancer) with no scanner-specific or population-specific tuning. We measured cancer detection (area under the ROC curve of the predicted cancer-tissue fraction), all-slide ISUP agreement of the deployed detect-then-grade pipeline (quadratic-weighted kappa, QWK), and grading agreement on pathologist-confirmed cancers, at the slide level and, because a case carries up to two slides, at the patient level. The reference reader was S.A.; thresholds and operating points were cross-validated leave-one-out; scanners were compared by paired within-biopsy bootstrap and confidence intervals confirmed by patient-cluster bootstrap.

**Results:** Detection was statistically equivalent across scanners (AUC 0.987 to 0.991; paired differences at most 0.003) and transferred to this non-Western cohort with no per-population tuning. At a 95% sensitivity operating point the deployed pipeline reached cross-validated all-slide QWK of 0.86, 0.81, and 0.86 (Grundium, Hamamatsu, Leica), matching the inter-pathologist ceiling of 0.81, against 0.23 to 0.62 for the ungated model. Grading of confirmed cancers was scanner dependent: the compact Grundium (0.63) did not differ from the clinical Leica (0.67; paired difference 0.04, 95% CI -0.03 to 0.11), while both exceeded Hamamatsu (0.44). Results held at the patient level, with grading somewhat lower for two scanners; the two slides of a case disagreed in grade in 43% of cases, and patient clustering did not widen the intervals.

**Conclusions:** Cancer-tissue fraction is a triage signal robust across scanner and transferable to an underrepresented population for detection, while grading is the scanner-sensitive step. Prostate grading models should be deployed as a detect-then-grade pipeline, with grading validated per device and confirmed on the local population.

## 1. Introduction

Prostate cancer is among the most common cancers in men worldwide [1]. Gleason pattern, summarized as the ISUP grade group, is the strongest tissue-based predictor of its outcome and directs treatment from active surveillance to radical therapy [2, 3]. Grading is also subjective. Inter-observer agreement between generalist pathologists is moderate, and standardization studies report substantial disagreement even among experts [4, 5]. This variability motivates computational support.

Deep-learning systems now grade prostate biopsies at a level comparable to pathologists [6]. Large studies from independent groups report high agreement with expert reference standards [7-9], and the PANDA challenge showed that many algorithms reach pathologist-level performance on curated data [10]. These results were obtained, with few exceptions, on images from one or a small number of high-end clinical scanners.

Scanner choice changes the image. Color response, focus, compression, and background differ by device, and this domain shift degrades models trained on a single source [11, 12]. Stain and color normalization and, more recently, pathology foundation models were introduced in part to reduce this sensitivity [13-16], though their robustness across scanners and populations is only beginning to be benchmarked [17]. The relevant question for deployment is not whether a model works on its development scanner, but whether one fixed model transfers across the scanners a laboratory or a low-resource site may use. Answering it requires the same physical slides imaged on several devices, which most datasets lack.

Population is a second, less studied source of shift. Grading systems and the foundation models beneath them are trained predominantly on North American and European material, yet they are proposed for use worldwide. Differences in ancestry, referral pattern, tissue fixation, and staining protocol between the development population and the deployment population can move the image and the label distribution together. Whether a model built on Western cohorts holds on a Middle Eastern population has, to our knowledge, been reported only in concurrent work on this same cohort [18], which we discuss below; the question remains almost entirely unexplored for automated prostate grading. PAR is drawn from Erbil, in the Kurdistan Region of Iraq, so it tests population and geographic transfer at the same time as scanner transfer.

The signal we test as a triage gate, the fraction of tissue occupied by cancer, is not itself new. Percentage of Gleason pattern 4 and percentage of tumour involvement are established biopsy quantities used in risk stratification [2, 3]. What is new here is its use as a scanner-robust gate in front of automated grading, and the measurement of how that gate behaves across devices.

The PAR dataset [19] allows the exploration of both shifts at once. Each of 339 prostate needle biopsies was digitized on three scanners spanning a clinical flagship, a common mid-tier device, and a low-cost compact scanner, and graded by three pathologists including a subspecialist. Concurrent with this work, the dataset originators evaluated a task-specific model and the UNI and Virchow2 foundation models on PAR, reporting that detection and grading generalized to this population and that model predictions were reproducible across the three scanners [18]. Our study is complementary in three respects: we evaluate a different, independently developed pipeline; we use the cancer-tissue fraction as an explicit triage gate ahead of grading; and where they characterized scanner effects through the model’s cross-scanner reproducibility, we add agreement with the pathologist measured on each scanner separately, for detection and for grading. Because the model we test was developed on Western data and PAR is a Middle Eastern cohort, applying it here without adaptation measures scanner and population transfer together. We separate two clinical tasks that a single ISUP number conflates, detecting cancer and grading it, and we report each at the slide level and at the patient level. All-slide agreement of the deployed pipeline governs workload in an all-comers setting; grading agreement on confirmed cancers isolates grading skill. We give particular attention to the compact scanner, the relevant device where a clinical digitizer is unaffordable, and to whether a Western-developed model holds on this population.

## 2. Materials and Methods

### 2.1. Dataset

PAR (EMBL-EBI BioImage Archive, accession S-BIAD2323, CC BY 4.0) comprises 339 haematoxylin-and-eosin prostate needle-biopsy slides from 185 patients in Erbil, Kurdistan Region of Iraq. Each slide was scanned on three devices: a Grundium Ocus40 (low-cost compact), a Hamamatsu NanoZoomer HT 2.0, and a Leica Aperio GT 450 DX (clinical reference), giving 1017 whole-slide images. Slide-level Gleason score and ISUP grade group were assigned independently by three readers: S.A. graded all 339 slides, H.M. graded 337, and a subspecialist uropathologist A.B. graded a 59-slide, ISUP-stratified subset. The dataset provides no consensus label and no pixel-level annotation. The 185 patients contributed 339 slides: 154 cases have two slides, labelled a and b, and 31 have one. The two slides of a case received different S.A. ISUP grades in 66 of 154 cases (43%), so slide-level grades are distinct labels rather than a single case grade recorded twice.

### 2.2. Model and inference

We used one fixed PathTools grading pipeline. Tissue is tiled at 20x into 224-pixel patches, and each patch is embedded into a 2560-dimensional vector by PathtoolsFM v1, a Vision-Transformer-Huge pathology foundation model [13, 16] (class token concatenated with the mean patch token). Grading then proceeds in stages: a two-stage cancer finder (version 14.1) detects cancerous regions with a binary classifier followed by a false-positive filter, and a region-level grader based on transformer multiple-instance learning with gated attention (TransMIL, version 13.1) assigns an ordinal Gleason pattern to each gland. A slide-level ISUP grade is formed by the needle-biopsy convention in which the primary pattern is the most extensive and the secondary is the highest-grade pattern present. The cancer-tissue fraction is the share of tissue assigned any Gleason pattern by this pipeline. All components were developed on prostate cohorts from North America and Europe [20] that do not include PAR, and the identical weights and thresholds were applied to all 1017 images with no scanner-specific or population-specific tuning and no color normalization. Inference ran on local RTX A6000 GPUs.

### 2.3. Evaluation and statistics

We evaluated three quantities per scanner. First, detection: the area under the ROC curve (AUC) of the cancer-tissue fraction as a score for clinically significant cancer, defined as ISUP grade 2 or higher, present in 168 of 339 biopsies (49.6%) under S.A. Second, all-slide grading agreement of the deployed detect-then-grade pipeline: slides with cancer-tissue fraction below an operating-point threshold are reported benign, the remainder keep the model grade, and the resulting ISUP is compared to the pathologist over all 339 slides. Third, grading agreement on pathologist-confirmed cancers. Agreement is the quadratic-weighted kappa (QWK) [21], interpreted against standard categories [22].

Pathologist S.A. is the primary reference because S.A. graded the full cohort. To guard against reproducing one reader’s idiosyncrasies, the all-slide result is also reported against H.M., and majority vote and highest grade are reported as sensitivity analyses. The detection operating-point threshold was fixed a priori at a target sensitivity (95%, 98%, or 99%) chosen on clinical grounds, not tuned to any agreement metric, and it was cross-validated leave-one-out: the threshold applied to each slide is estimated from the other 338 slides and never sees its own label. Confidence intervals (95%) come from 2000 bootstrap resamples of slides. Because every biopsy is imaged on all three scanners, scanners are compared by paired within-biopsy bootstrap of the metric difference, which is exact for this matched design and is reported in preference to comparing separate per-scanner intervals.

Each quantity is computed at two units. The slide is the primary unit, because one physical slide is imaged on all three devices and so isolates the scanner effect on identical tissue. The patient (case) is the clinically reported unit; a case grade is the highest ISUP across its slides and a case detection score is the highest cancer-tissue fraction across its slides, following the needle-biopsy worst-grade convention. Because slides cluster within patients, every slide-level confidence interval was also recomputed by resampling patients rather than slides.

## 3. Results

Figure 1 orients the analysis with one illustrative biopsy imaged on the three scanners. The same cores are recognizable on every device, color and background differ visibly, and the model localizes and grades the tumor consistently. This single concordant case is shown for illustration only; the quantitative results follow.

**Figure 1.**
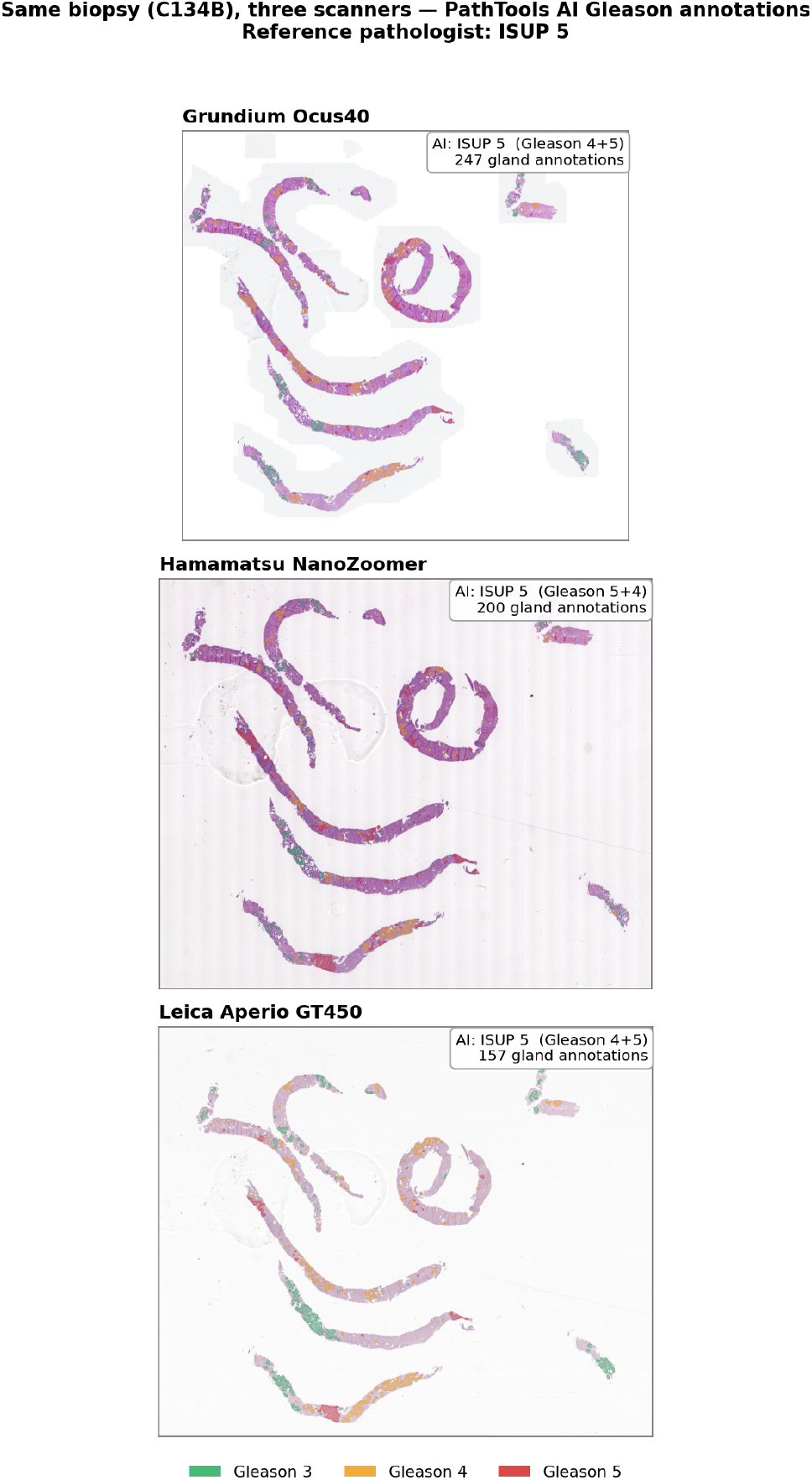
One illustrative biopsy (C134B) imaged on three scanners with PathTools gland-level Gleason annotations (green, pattern 3; amber, pattern 4; red, pattern 5). Reference ISUP 5; the model returned ISUP 5 on all three scanners. Shown as a single example, not as representative performance.

### 3.1. Reference-standard reliability

The two full-cohort readers agreed at QWK 0.81 (95% CI 0.76 to 0.85) yet differed in grade distribution (Table 1): S.A. assigned more intermediate grades, H.M. more benign and more ISUP 5. Against the subspecialist uropathologist on the 59-slide overlap, S.A. reached QWK 0.84 (95% CI 0.70 to 0.93). S.A. is therefore a defensible full-cohort reference. These figures also set the human ceiling against which the model is measured.

**Table 1.** Slide-level ISUP distribution by reader. The full-cohort readers diverge, which motivates the reference-rule sensitivity analysis in Table 3.

| ISUP | S.A. (n=339) | H.M. (n=337) | A.B. (n=59) |
| --- | --- | --- | --- |
| 0 (benign) | 164 | 209 | 11 |
| 1 | 7 | 2 | 5 |
| 2 | 38 | 7 | 13 |
| 3 | 59 | 8 | 3 |
| 4 | 30 | 25 | 6 |
| 5 | 41 | 86 | 21 |

### 3.2. Detection of cancer is scanner-robust

The predicted cancer-tissue fraction separated significant cancer from benign nearly perfectly on every scanner (Figure 2): AUC 0.987 (95% CI 0.978 to 0.994) for the compact Grundium, 0.987 (0.977 to 0.995) for Hamamatsu, and 0.991 (0.981 to 0.997) for Leica. On the matched biopsies, every paired between-scanner AUC difference was within 0.01 with a confidence interval spanning zero (Leica minus Grundium 0.003, 95% CI -0.004 to 0.010; Grundium minus Hamamatsu 0.000, -0.005 to 0.006), so detection showed no between-scanner difference within a 0.01 AUC margin. In a rule-out workflow with leave-one-out thresholds (Table 2), specificity at 98% sensitivity was 87 to 92% and 45 to 48% of biopsies were routed to auto-benign. We note that negative predictive value and cleared fraction depend on the cohort prevalence of 49.6%, which is enriched relative to a screening population and is not directly transferable to one.

**Table 2.** Rule-out operating points for significant-cancer triage, leave-one-out cross-validated (S.A. reference). NPV is the probability that an auto-cleared biopsy is truly benign and ‘cleared’ is the share of all biopsies routed to auto-benign; both depend on the cohort cancer prevalence of 49.6% and are cohort-specific, not screening-population estimates. Grundium and Leica coincide at 95% sensitivity because their leave-one-out thresholds yield identical confusion counts on this cohort, not through any data-handling error.

| Scanner | Target sensitivity | Specificity | NPV | Cleared |
| --- | --- | --- | --- | --- |
| Grundium | 95% | 95.3% | 94.8% | 50.7% |
|  | 98% | 86.5% | 97.4% | 44.8% |
|  | 99% | 77.8% | 98.5% | 39.8% |
| Hamamatsu | 95% | 92.4% | 94.6% | 49.3% |
|  | 98% | 88.9% | 97.4% | 46.0% |
|  | 99% | 82.5% | 98.6% | 42.2% |
| Leica | 95% | 95.3% | 94.8% | 50.7% |
|  | 98% | 91.8% | 97.5% | 47.5% |
|  | 99% | 88.3% | 98.7% | 45.1% |

**Figure 2.**
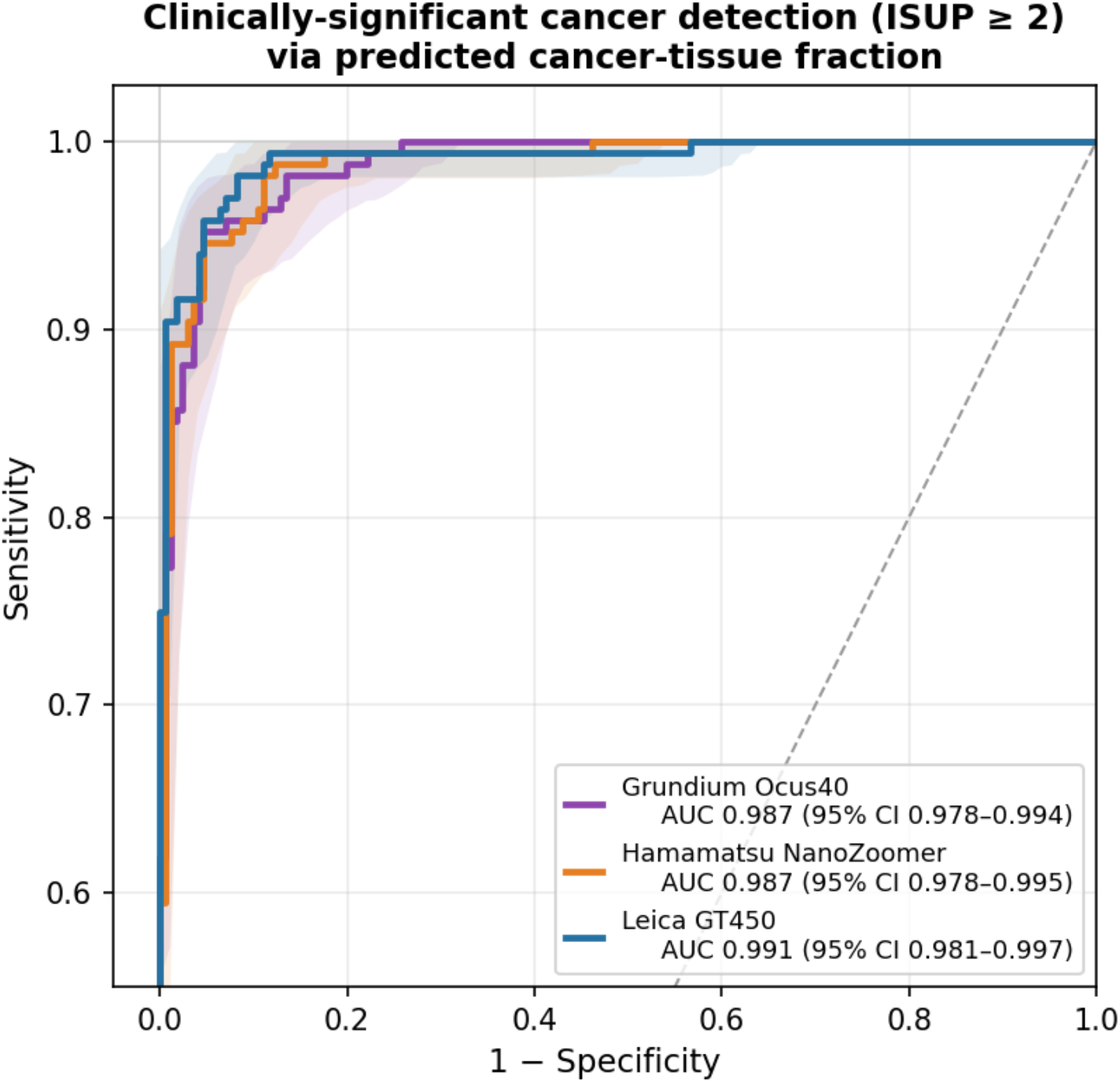
Detection of clinically significant cancer (ISUP grade 2 or higher) via the predicted cancer-tissue fraction. Shaded regions are bootstrap 95% CI bands; all three AUC intervals overlap.

### 3.3. The deployed pipeline recovers near-ceiling all-slide agreement

The ungated model agreed poorly with the pathologist across all comers, and unevenly by scanner: all-slide QWK 0.23 (Grundium), 0.42 (Hamamatsu), and 0.62 (Leica). The cause is benign over-call, worst on the compact scanner, where sparse spurious high-grade patches inflate the ISUP of benign slides. Gating on the cancer-tissue fraction removes these over-calls. With the threshold cross-validated leave-one-out at a 95% sensitivity operating point, all-slide QWK reached 0.86 (95% CI 0.81 to 0.90) for Grundium, 0.81 (0.75 to 0.85) for Hamamatsu, and 0.86 (0.82 to 0.90) for Leica, matching the interpathologist ceiling of 0.81 on all three (Table 3).

**Table 3.**
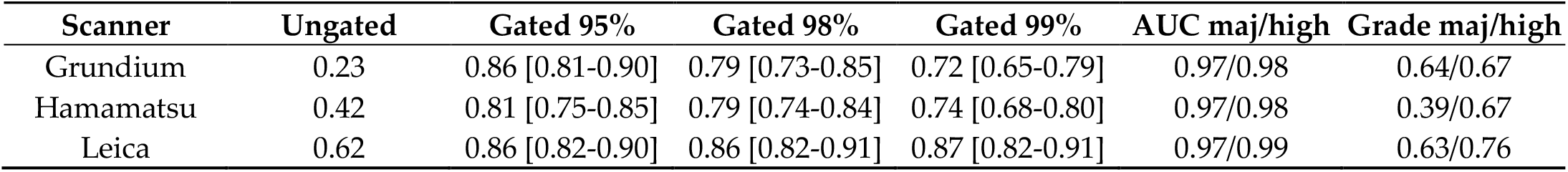
Deployed detect-then-grade pipeline. All-slide QWK against S.A. over all 339 slides (95% CI in brackets), with the detection gate cross-validated leave-one-out at three target sensitivities. ‘Ungated’ keeps every model grade. Final columns give detection AUC and cancer-grading QWK under majority-vote and highest-grade references as sensitivity analyses.

| Scanner | Ungated | Gated 95% | Gated 98% | Gated 99% | AUC maj/high | Grade maj/high |
| --- | --- | --- | --- | --- | --- | --- |
| Grundium | 0.23 | 0.86 [0.81-0.90] | 0.79 [0.73-0.85] | 0.72 [0.65-0.79] | 0.97/0.98 | 0.64/0.67 |
| Hamamatsu | 0.42 | 0.81 [0.75-0.85] | 0.79 [0.74-0.84] | 0.74 [0.68-0.80] | 0.97/0.98 | 0.39/0.67 |
| Leica | 0.62 | 0.86 [0.82-0.90] | 0.86 [0.82-0.91] | 0.87 [0.82-0.91] | 0.97/0.99 | 0.63/0.76 |

The operating point trades sensitivity against agreement. At 98% sensitivity the two scanners with more benign over-call fell to 0.79 (0.73 to 0.85) and 0.79 (0.74 to 0.84), each below the 0.81 point estimate but within its confidence interval, while Leica held at 0.86. Two controls argue against an artifact of threshold selection. First, leave-one-out estimation, in which each slide’s threshold is set on the other 338, changed the values by no more than 0.006 from the in-cohort estimate, so the near-match is not driven by a threshold fitted to the same slides. Second, the same gated pipeline evaluated against the second reader H.M., rather than the reference S.A., gave similar agreement (0.82, 0.74, and 0.86 at 98% sensitivity), so the result is not an artifact of scoring the model against the single reader whose reads it most resembles.

### 3.4. Grading of confirmed cancers is the scanner-sensitive step

On pathologist-confirmed cancers, grading agreement was 0.63 (95% CI 0.54 to 0.71) for Grundium, 0.44 (0.34 to 0.53) for Hamamatsu, and 0.67 (0.58 to 0.74) for Leica, all below the inter-pathologist ceiling of 0.81 (the grading intervals and the ceiling interval do not overlap). On the matched biopsies, Grundium and Leica did not differ (paired difference 0.04, 95% CI -0.03 to 0.11), so the compact scanner was not detectably worse than the clinical one, though the study is not powered for equivalence. Both exceeded Hamamatsu by margins whose intervals excluded zero (Leica minus Hamamatsu 0.23, 0.15 to 0.31; Grundium minus Hamamatsu 0.19, 0.11 to 0.27).

The Hamamatsu deficit is directional, not random. Its confusion matrix (Figure 3) shows intermediate-grade cancers collapsing onto ISUP 5. Consistent with an upward shift, Hamamatsu grading rose from 0.44 against S.A. to 0.67 against a highest-grade reference (Table 3). The low ungated all-slide agreement on Grundium (Section 3.3) therefore reflects benign over-call rather than poor grading of true cancers, whereas the Hamamatsu deficit is a grading effect.

**Figure 3.**
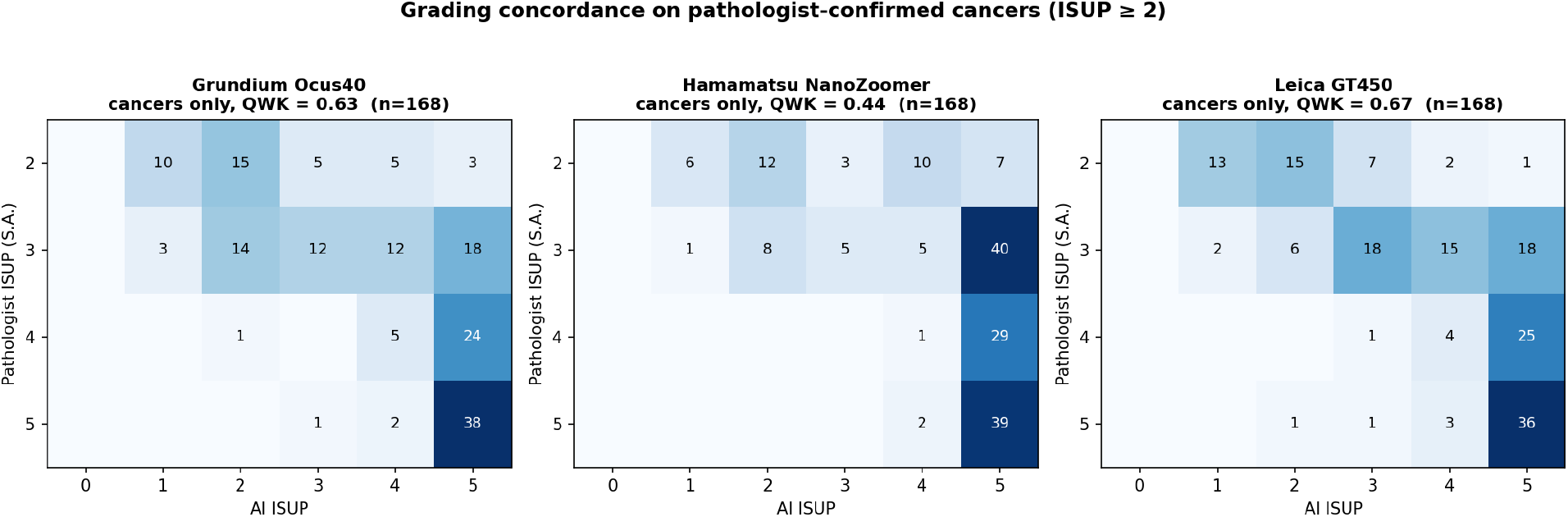
Grading concordance (model versus S.A.) on pathologist-confirmed cancers. Grundium and Leica track the diagonal; Hamamatsu over-assigns ISUP 5.

### 3.5. Slide-level findings hold at the patient level

Grades were assigned per slide, and the two slides of a case disagreed in 43% of cases (Section 2.1), so we repeated the analysis at the patient level under the worst-grade convention (Table 4). Detection was essentially unchanged (case-level AUC 0.985, 0.984, and 0.987). Grading agreement on confirmed cancer cases was 0.54, 0.33, and 0.68 for Grundium, Hamamatsu, and Leica: lower than the slide level for the two scanners that already lagged and unchanged for Leica, while the scanner ordering held. Patient clustering did not widen the intervals; the Leica grading interval was [0.58, 0.74] resampling slides and [0.57, 0.74] resampling patients. The clinically reported unit is therefore consistent with the slide-level headline for detection, and a little more demanding for grading.

**Table 4.**
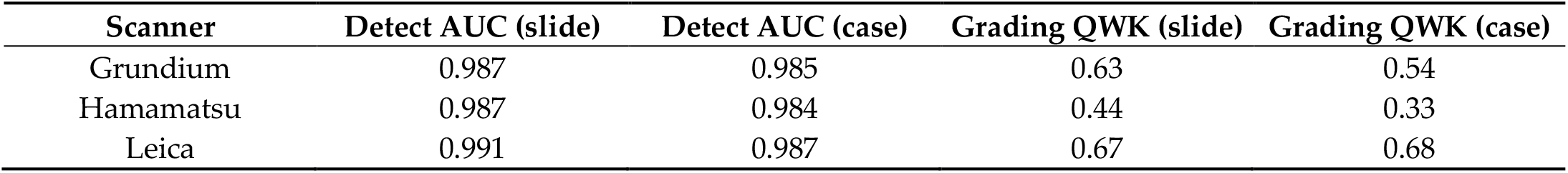
Slide-level versus patient-level agreement (S.A. reference). Patient level aggregates a case’s slides by the worst-grade convention. Detection is level-invariant; case-level grading is lower for Grundium and Hamamatsu, and the scanner ordering is preserved.

| Scanner | Detect AUC (slide) | Detect AUC (case) | Grading QWK (slide) | Grading QWK (case) |
| --- | --- | --- | --- | --- |
| Grundium | 0.987 | 0.985 | 0.63 | 0.54 |
| Hamamatsu | 0.987 | 0.984 | 0.44 | 0.33 |
| Leica | 0.991 | 0.987 | 0.67 | 0.68 |

## 4. Discussion

One fixed model transferred its cancer-detection signal across three scanners but not its raw grade calibration. Paired analysis showed no between-scanner difference in detection within a 0.01 AUC margin, including a low-cost compact scanner, while ungated all-slide agreement ranged from 0.23 to 0.62 and was worst on the compact device. The gap is explained by benign over-call: the model assigns sparse high-grade patches to benign tissue, which inflates ISUP without changing the underlying benign-versus-cancer signal. Gating on cancer-tissue fraction closed the gap and lifted cross-validated all-slide agreement to the inter-pathologist ceiling at a 95% sensitivity operating point on all three scanners, with a modest and expected decline as the operating point is pushed to higher sensitivity.

The practical consequence is a detect-then-grade workflow. A detection stage auto-clears a large fraction of benign biopsies and needs no per-scanner recalibration. The magnitude of that saving is prevalence-dependent: in this cohort, which is roughly half clinically significant cancer, 45 to 48% of biopsies were cleared at 98% sensitivity, and the negative predictive value and cleared fraction would both rise in a lower-prevalence screening setting and should be re-estimated there. Grading of the remaining cancers is where scanner choice matters and should be validated per device. The mid-tier Hamamatsu showed a directional Gleason-5 bias that is a candidate for stain normalization or grade recalibration, although part of that direction may be a reference effect, since the second reader H.M. also assigned ISUP 5 more often than S.A. (86 versus 41 slides). The compact scanner graded cancers with no detectable difference from the clinical reference.

The compact-scanner result has direct relevance to low-resource and non-digitized settings. A device costing a fraction of a clinical digitizer supported both scanner-robust triage and cancer grading comparable to the flagship scanner. This supports pairing inexpensive hardware with a validated triage model where a full digital-pathology deployment is not feasible.

A concurrent study by the dataset originators [18] evaluated a task-specific model and the UNI and Virchow2 foundation models on this cohort. That study found detection and grading concordance with pathologists within the inter-observer range, and high cross-scanner reproducibility (quadratic-weighted kappa above 0.90) when comparing each model’s predictions across scanner pairs. Our results are consistent on detection and on population transfer, and extend that work in a specific way. Cross-scanner reproducibility assesses whether a model returns the same grade on the same slide across devices; per-scanner agreement with the pathologist asks whether that grade is correct on each device. These can diverge, and for the pipeline tested here they do: grading agreement with the reference was scanner-dependent, with a directional Gleason-5 shift on the Hamamatsu, even though detection was not. The two studies therefore point the same way on detection and, taken together, refine the grading message: whether grading transfers across scanners is model-dependent, which is why we argue for validating the grading stage per device rather than assuming the reproducibility of any one model.

The model was built on North American and European material and applied to a cohort from Erbil with no adaptation. Its detection of clinically significant cancer transferred intact, evidence that the benign-versus-cancer signal of a Western-developed model can carry to a population underrepresented in its training data. In companion work, the same automated grading approach stratifies disease-free interval from a single out-of-distribution slide [23].

We frame this as external validation, not as a measurement of a population effect. PAR has no Western arm imaged on the same scanners, so we cannot attribute any part of the residual grading gap to population rather than device; the two change together relative to the development data. Differences plausibly present between an Iraqi cohort and Western development cohorts, in ancestry, referral pattern, fixation, and staining, could shift both the images and the labels, and they would be absorbed into, not separated from, the scanner-dependent grading result. What we can state is bounded: detection generalized across device and population, grading did so only in part, and a single Middle Eastern site does not support wider geographic claims. The practical reading is cautious optimism. A Western-developed triage model carried its detection value to this population and to inexpensive hardware, but grading on a new population and scanner should be confirmed locally before autonomous use, and assisted reading carries its own anchoring risks that must be managed [24].

Reassuringly, the picture did not change at the patient level, the unit that reaches a clinical report. Detection was level-invariant and the scanner ordering held, though grading fell somewhat at the case level for the two scanners that already lagged. Because the two slides of a case disagree in grade almost half the time, this patient-level check confirms that the slide-level headline is not an artifact of counting correlated slides, a conclusion the patient-cluster bootstrap supports directly.

Our cancer-restricted grading agreement (0.44 to 0.67) sits in the same range as the malignant-only concordance reported for other models on this cohort (0.62 to 0.66 [18]) and below the highest single-scanner reports [7, 9, 10]. On the same data, the pipeline tested here detects clinically significant cancer more accurately (AUC 0.987 to 0.991 vs 0.916 to 0.966 [18]) but grades the Hamamatsu less well. This is expected. Those studies fine-tune or select on their target distribution, compare against consensus panels, and evaluate on one scanner. We applied a fixed model, without any adaptation, against a single-reader reference, across three scanners. The comparison that matters here is internal and controlled: the same model on the same slides differs by scanner, and by task.

We report both all-slide and cancer-restricted agreement deliberately. As a general property, when benign cases predominate the all-slide QWK is dominated by benign-versus-cancer separation and can be high even when grading skill among cancers is lower; the present cohort is not benign-heavy, at 49.6% cancer, but a real screening population would be, which underscores the importance of the all-comers number. A screening service cares about the all-slide number, and a grading laboratory about the cancer-restricted one.

### 4.1. Limitations

This study has several limitations of data and reference standard. It draws on a single Middle Eastern site, Erbil, and a model developed on Western cohorts, which makes PAR a useful population-transfer test but leaves geographic and population generalization beyond one Middle Eastern cohort, and the separation of population from scanner effects, unproven. The reference standard is single-reader-dominant: no consensus label or pixel-level annotation exists for PAR, the two full-cohort readers agree only moderately, and scoring the model against S.A. while comparing to an S.A.-based ceiling shares a common reader. We mitigate the last point by also reporting agreement against H.M., which was similar, but it is not fully removed. The cohort is also enriched, at 49.6% clinically significant cancer, so the negative predictive value and cleared fraction are prevalence-dependent and are not screening-population estimates.

Limitations of design and analysis follow. Several operating points and three reference rules were examined; the headline operating point was fixed a priori on clinical grounds and thresholds were cross-validated leave-one-out, which moved values by no more than 0.006, but the multiplicity should be kept in mind and a prospective threshold confirmed before clinical use. The study is not powered for formal equivalence, so a finding of no detectable difference between the compact and clinical scanners is not proof of equivalence. One fixed inference model was tested, so the findings characterize this model on PAR rather than prostate AI in general. Finally, the design is retrospective and does not measure effects on patient outcomes or reader behavior.

## 5. Conclusion

Cancer-tissue fraction is a triage signal that held across three scanners and transferred, for detection, to a Middle Eastern population outside the model’s Western development data. A detect-then-grade pipeline built on it recovered all-slide agreement close to the inter-pathologist ceiling on every scanner, including a low-cost compact device, at both the slide and the patient level, whereas the ungated model did not. Grading of confirmed cancers, not detection, was the scanner-sensitive step. These results support deploying prostate grading models as a two-stage system, validating the grading stage on each device, and confirming performance on the local population, and they indicate that inexpensive compact scanners are worth further study within such a system.

## Data Availability

All data produced in the present study are available upon reasonable request to the authors.

## Author Contributions

Conceptualization, D.DC; methodology, D.DC; software, D.DC; validation, J.L.E, A.P, J.S, D.DC ; formal analysis, D.DC ; investigation, J.L.E, D.DC; resources, D.DC; data curation, D.DC; writing—original draft preparation, J.L.E; writing—review and editing, A.P, J.S, D.DC; visualization, D.DC; supervision, D.DC; project administration, D.DC; funding acquisition, D.DC. All authors have read and agreed to the published version of the manuscript.

## Institutional Review Board Statement

Ethical review and approval were not required for this study due to its nature as an analysis of a public, de-identified dataset (EMBL-EBI BioImage Archive S-BIAD2323).

## Informed Consent Statement

Not applicable

## Data Availability Statement

The PAR dataset is available at EMBL-EBI BioImage Archive, accession S-BIAD2323 (CC BY 4.0). Analysis code is available from the authors on reasonable request.

## Acknowledgments

AI (Claude Opus 5) was used to as writing aid of this article.

## Abbreviations

The following abbreviations are used in this manuscript:

AI: Artificial Intelligence
AUC: Area Under the Curve
CC BY 4.0: Creative Commons Attribution 4.0 Internatinoal License CI Confidence Interval
EMBL-EBI: European Molecular Biology Laboratory-European Bioinformatics Institute
GPU: Graphics Processing Unit
ISUP: International Society of Urological Pathology
NPV: Negative Predictive Value
PANDA: Prostate cANcer graDe Assessment
QWK: Quadratic-Weighted Kappa
ROC: Receiver Operating Characteristic
TransMIL: Transformer Multiple Instance Learning

## Disclaimer/Publisher’s Note

The statements, opinions and data contained in all publications are solely those of the individual author(s) and contributor(s) and not of MDPI and/or the editor(s). MDPI and/or the editor(s) disclaim responsibility for any injury to people or property resulting from any ideas, methods, instructions or products referred to in the content.

## References

1. Sung, H., et al., Global cancer statistics 2020: GLOBOCAN estimates of incidence and mortality worldwide for 36 cancers in 185 countries. CA: a cancer journal for clinicians, 2021. 71(3): p. 209–249.

2. Epstein, J.I., et al., The 2014 International Society of Urological Pathology (ISUP) consensus conference on Gleason grading of prostatic carcinoma: definition of grading patterns and proposal for a new grading system. The American journal of surgical pathology, 2016. 40(2): p. 244–252.

3. Epstein, J.I., et al., A contemporary prostate cancer grading system: a validated alternative to the Gleason score. European urology, 2016. 69(3): p. 428–435.

4. Egevad, L., et al., Standardization of Gleason grading among 337 European pathologists. Histopathology, 2013. 62(2): p. 247–256.

5. Ozkan, T.A., et al., Interobserver variability in Gleason histological grading of prostate cancer. Scandinavian journal of urology, 2016. 50(6): p. 420–424.

6. Frewing, A., et al., Don’t fear the artificial intelligence: a systematic review of machine learning for prostate cancer detection in pathology. Archives of Pathology & Laboratory Medicine, 2024. 148(5): p. 603–612.

7. Bulten, W., et al., Automated deep-learning system for Gleason grading of prostate cancer using biopsies: a diagnostic study. The Lancet Oncology, 2020. 21(2): p. 233–241.

8. Nagpal, K., et al., Development and validation of a deep learning algorithm for improving Gleason scoring of prostate cancer. NPJ digital medicine, 2019. 2(1): p. 48.

9. Ström, P., et al., Artificial intelligence for diagnosis and grading of prostate cancer in biopsies: a population-based, diagnostic study. The lancet oncology, 2020. 21(2): p. 222–232.

10. Bulten, W., et al., Artificial intelligence for diagnosis and Gleason grading of prostate cancer: the PANDA challenge. Nature medicine, 2022. 28(1): p. 154–163.

11. Campanella, G., et al., Clinical-grade computational pathology using weakly supervised deep learning on whole slide images. Nature medicine, 2019. 25(8): p. 1301–1309.

12. Van der Laak, J., G. Litjens, and F. Ciompi, Deep learning in histopathology: the path to the clinic. Nature medicine, 2021. 27(5): p. 775–784.

13. Chen, R.J., et al., Towards a general-purpose foundation model for computational pathology. Nature medicine, 2024. 30(3): p. 850–862.

14. Macenko, M., et al. A method for normalizing histology slides for quantitative analysis. in 2009 IEEE international symposium on biomedical imaging: from nano to macro. 2009. IEEE.

15. Tellez, D., et al., Quantifying the effects of data augmentation and stain color normalization in convolutional neural networks for computational pathology. Medical image analysis, 2019. 58: p. 101544.

16. Vorontsov, E., et al., A foundation model for clinical-grade computational pathology and rare cancers detection. Nature medicine, 2024. 30(10): p. 2924–2935.

17. Ebbert, J.L. and D. Della Corte, PANDA-PLUS-Bench: A Clinical Benchmark for Evaluating the Robustness of AI Foundation Models in Prostate Cancer Diagnosis. AI in Medicine, 2026. 1(2): p. 14.

18. Ali, P.J.M., et al., Validation of Diagnostic Artificial Intelligence Models for Prostate Pathology in a Middle Eastern Cohort. arXiv preprint arXiv:2512.17499, 2025.

19. Muhammad Ali, P.J., et al., The PAR dataset: Prostate biopsy whole slide images from an underrepresented Middle Eastern population. Scientific Data, 2026. 13(1): p. 1061.

20. Hopson, S., et al., PANDA-PLUS: Improved dataset of prostate whole slide images from PANDA Challenge with pixel-level expert annotations. Journal of Pathology Informatics, 2026. 20: p. 100540.

21. Cohen, J., Weighted kappa: Nominal scale agreement provision for scaled disagreement or partial credit. Psychological bulletin, 1968. 70(4): p. 213.

22. Landis, J.R. and G.G. Koch, The measurement of observer agreement for categorical data. biometrics, 1977: p. 159–174.

23. Ebbert, J.L., et al., An Automated, Pathologist-free Gleason Grade Stratifies Disease-free Interval Comparably to Expert Grading from a Single Out-of-distribution Slide. medRxiv, 2026: p. 2026.06. 22.26356247.

24. Perry, A., et al., Anchoring Bias in AI-Assisted Gleason Grading Across the Expertise Spectrum: A Pre-Registered, Within-Subjects Study. 2026.

